# Simultaneous quantification of drugs in whole blood by ultra-performance liquid chromatography–tandem mass spectrometry using solid-phase mini-cartridges (Smart-SPE) for sample preparation

**DOI:** 10.64898/2026.01.12.26343926

**Authors:** Takayoshi Suzuki, Naoto Judai, Miyu Ishihara, Kiri Kajimoto

## Abstract

**Objective:** Drug poisoning cases due to overdoses of over-the-counter (OTC) medications are increasing, and comprehensive measurement of blood drug concentrations, including OTC drugs, is important in emergency medicine and forensic science. In this study, we developed a simultaneous quantification method for blood drug levels using LC–MS/MS with solid-phase mini-cartridge SmartSPE for sample preparation.

**Methods:** The target analytes were acetaminophen, caffeine, flunitrazepam, 7-aminoflunitrazepam, risperidone, and phenobarbital. Internal standards (IS) used were acetaminophen-*d*_4_, caffeine-*d*_9_, diazepam-*d*_5_, and phenobarbital-*d*_5_. For solid-phase extraction, three types of Smart-SPE columns (AiSTI Science Co., Ltd.) were employed. 100 µL of blank whole blood samples were subjected to protein precipitation using methanol and acetonitrile, and the supernatant obtained after centrifugation was processed using Smart-SPE for sample cleanup. Chromatographic separation was performed on a CAPCELL PAK INERT ADME-HR column (Osaka Soda), and detection was carried out in both positive and negative ESI modes.

**Results:** Linearity was observed for all drugs across the therapeutic to coma-death concentration ranges, with correlation coefficients exceeding 0.99 in all cases. Intra-day accuracy ranged from 98.06% to 110.87%, with precision between 0.19% and 17.37%. Inter-day accuracy ranged from 97.82% to 112.35%, with precision between 0.53% and 9.18%. Recovery rates varied from 72.9% to 95.1%, and matrix effects ranged from 91.2% to 113.0%.

**Discussion:** The validation results demonstrated satisfactory performance for all analytes, suggesting that this simultaneous quantification method may serve as a reliable analytical approach. Further investigations into sample stability are planned, and if analyses of actual specimens confirm its applicability, the method can be reported as a novel approach for simultaneous quantification of blood drug levels using a new sample pretreatment technique.

## Introduction

The incidence of drug poisoning has been increasing in recent years. Notably, in adolescents, over 50% of all cases are attributable to over-the-counter (OTC) medications, representing a significant public health concern. In contrast, the proportion of poisoning cases caused by psychoactive drugs has remained relatively stable over the past decade, accounting for approximately 30% of all cases. Together with OTC medications, these substances are involved in roughly 50% of all poisoning incidents^1)^.

Common OTC medications involved in poisoning include those containing caffeine or acetaminophen, which are frequently reported in overdose cases. These drugs are primarily obtained through in-store purchases at drugstores^2)^, making them relatively accessible to adolescents and contributing to an increasing incidence of poisoning among young people.

Among psychoactive drugs, benzodiazepine anxiolytics such as flunitrazepam and etizolam are the most frequently involved in poisoning cases. Other reported drugs include barbiturates (e.g., phenobarbital), atypical antipsychotics (e.g., risperidone and paliperidone), and hypnotics such as zolpidem^3-7)^. Poisoning with psychoactive drugs is often associated with high severity and urgency in emergency medical care, necessitating prompt and appropriate treatment^8)^. Because management strategies vary depending on the causative agent, identification of the specific drug and measurement of its blood concentration are essential.

In forensic medicine, deaths due to overdose of psychoactive drugs or caffeine have been reported^9-12)^. Therefore, comprehensive measurement of blood drug concentrations, including over-the-counter medications, is essential for accurately determining the cause of death.

For drug concentration analysis, biological samples such as whole blood or serum are commonly used. Liquid chromatography-tandem mass spectrometry (LC–MS/MS) is frequently employed due to its relatively short analysis time^13)^. When analyzing drugs in biological samples, a typical procedure involves deproteinization and extraction using organic solvents, followed by concentration of the extract to prepare the analytical sample. However, deproteinization with organic solvents alone may be insufficient to remove phospholipids, which can contaminate the LC–MS/MS ion source, reduce analytical sensitivity, and accelerate deterioration of the separation column^14)^.

To address this issue, solid-phase extraction (SPE) columns specifically designed to remove phospholipids are sometimes incorporated into the sample preparation process. In general, SPE works by retaining target compounds on a solid phase, washing away impurities, and subsequently eluting the analytes. This approach takes advantage of the hydrophobic nature of many drugs, allowing aqueous washes to remove impurities while retaining the target compounds, which are then selectively eluted with organic solvents.

However, because caffeine and acetaminophen are highly polar, they may be eluted during the aqueous washing step, potentially reducing analytical sensitivity. In this study, we employed a solid-phase mini cartridge, Smart-SPE, which does not require washing steps, to retain impurities on the solid phase while selectively eluting the target compounds. This approach aimed to enhance phospholipid removal and improve analytical sensitivity. The target analytes included OTC drugs such as caffeine and acetaminophen, as well as psychoactive drugs including flunitrazepam, phenobarbital, and risperidone. Additionally, since flunitrazepam is rapidly metabolized in vivo to 7-aminoflunitrazepam, this metabolite was also included as a target analyte.

## Materials and Methods

### Chemicals and Solid-Phase Columns

Acetaminophen, caffeine, flunitrazepam, 7-aminoflunitrazepam, and diazepam-*d*_5_ were purchased from Sigma-Aldrich (St. Louis, Mo, USA). Phenobarbital and risperidone were obtained from Tokyo Chemical Industry Co., Ltd. (Tokyo, Japan). Phenobarbital-*d*_5_ was purchased from Merck KGaA (Darmstadt, Germany), acetaminophen-*d*_4_ from Cayman Chemical Company (Ann Arbor, MI, USA), and caffeine-*d*_9_ from CDN Isotopes Inc. (Pointe-Claire, Quebec, Canada). The chemical structures of each substance are shown in Figure 1.

**Figure. 1.**
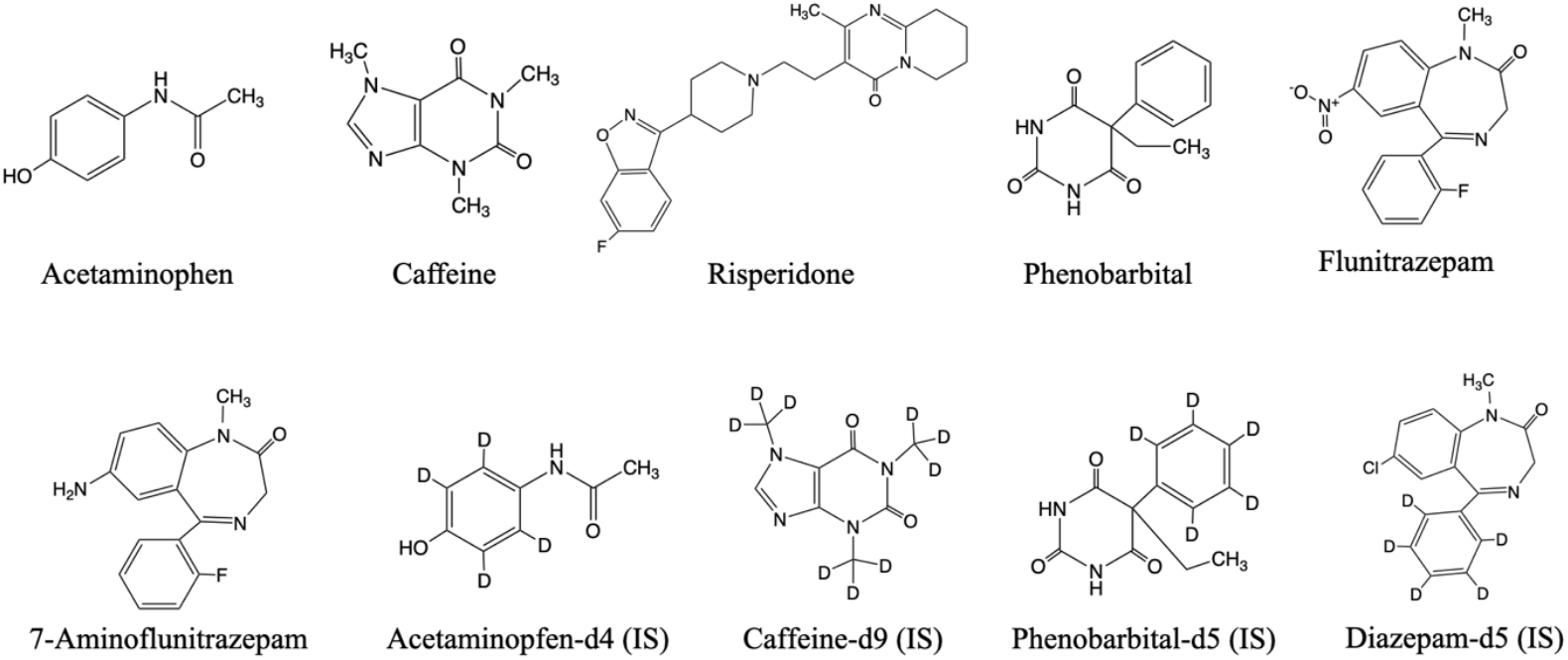
Chemical structures of the analytes and internal standards.

Flunitrazepam, 7-aminoflunitrazepam, risperidone, and diazepam-*d*_5_ were dissolved in methanol at a concentration of 10 μg/mL, phenobarbital-*d*_5_ at 20 μg/mL, caffeine-*d*_9_ at 100 μg/mL, and acetaminophen-*d*_4_ at 200 μg/mL. Caffeine, acetaminophen, and phenobarbital were prepared in methanol at concentrations of 1 mg/mL, 5 mg/mL, and 10 mg/mL, respectively. All solutions were stored at –30°C until use.

LC-MS/MS grade acetonitrile, methanol, and formic acid were purchased from Fujifilm Wako Pure Chemicals (Osaka, Japan). Purified water was obtained from the Direct-Q UV 3 system (Millipore, Burlington, MA, USA). Human whole blood was purchased from Tennessee Blood Services (Memphis, TN, USA).

For solid-phase extraction, three types of cartridges were used: Smart-SPE C18-50, C18-30, and PSA-30, all from AiSTI Science Co., Ltd. (Wakayama, Japan).

### Evaluation of Phospholipids Removal Effect of Solid-Phase Mini Cartridges

To 100 µL of whole blood, 170 µL of methanol containing internal standards (IS) and 300 µL of acetonitrile were added, mixed thoroughly, and centrifuged at 20,000 × g for 10 minutes to obtain the supernatant. Three different methods were then applied to the supernatant to prepare the measurement samples, and a comparative study was performed. The flowchart is shown in Figure 2.

**Figure. 2.**
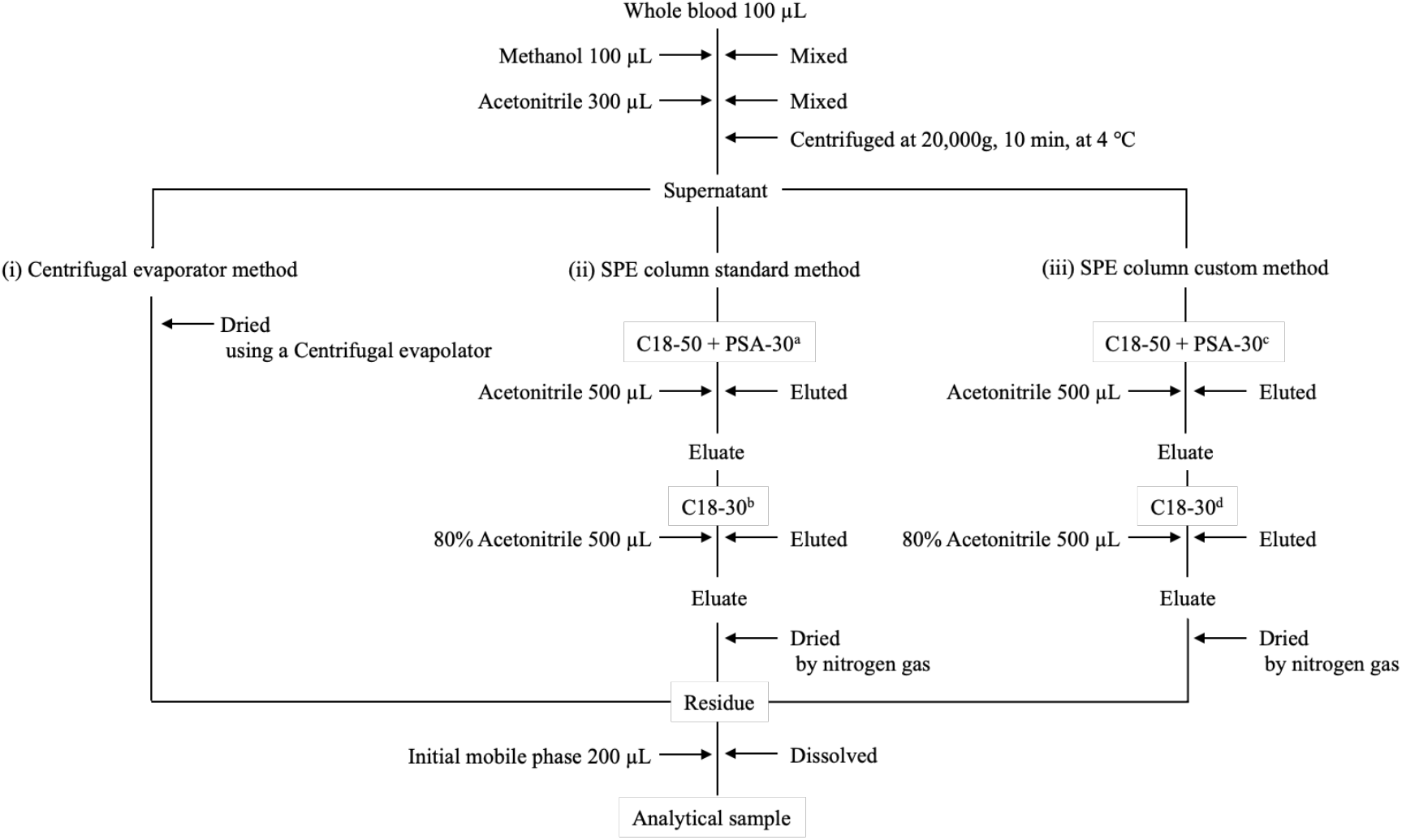
Process of the method for evaluating phospholipid removal efficiency using solid-phase mini cartridges ^a^The column was conditioned by eluting 2 mL of 80% acetonitrile, followed by 2 mL of 100% acetonitrile. ^b^The column was conditioned by eluting 2 mL of 100% acetonitrile, followed by 2 mL of 80% acetonitrile. ^c^The column was conditioned by eluting 2 mL of 100% acetonitrile, followed by 2 mL of 60% acetonitrile/ 20% methanol/ 20% H_2_0. ^d^The column was conditioned by eluting 2 mL of 100% acetonitrile, followed by 2 mL of 80% acetonitrile’ 10% methanol/ 10% H_2_0.

(i) Centrifugal Evaporator Method: The supernatant was dried using a centrifugal evaporator, then dissolved in 200 µL of the initial mobile phase.

(ii) SPE column standard method: A combined C18-50/PSA-30 column was activated with 2 mL of 80% acetonitrile, followed by 2 mL of 100% acetonitrile. The supernatant was then passed through the column, and the eluate was collected with 500 µL of 100% acetonitrile. For the C18-30 column, activation was performed with 2 mL of 100% acetonitrile followed by 2 mL of 80% acetonitrile. After passing the supernatant through the column, the eluate was collected with 500 µL of 80% acetonitrile. The collected eluates were dried under a stream of nitrogen and reconstituted in 200 µL of the initial mobile phase.

(ii) SPE column custom method: A combined column of C18-50 and PSA-30 was conditioned with 2 mL of 100% acetonitrile and 2 mL of a mixture of 60% acetonitrile, 20% methanol and 20% pure water. After passing the supernatant through the column, elution was performed with 500 µL of 100% acetonitrile. For the C18-30 column, it was conditioned with 2 mL of 100% acetonitrile and 2 mL of a mixture of 80% acetonitrile, 10% methanol and 10% pure water. After passing the sample through, the eluted solution was collected and then eluted with 500 µL of 80% acetonitrile.

The obtained solution was dried under nitrogen gas and then dissolved in 200 µL of the initial mobile phase.

### Drug Quantification Method

Based on the results from *Evaluation of Phospholipids Removal Effect of Solid-Phase Mini Cartridges*, 100 µL of whole blood was mixed with 170 µL of methanol containing internal standards (IS) and 300 µL of acetonitrile, followed by vortexing. The mixture was centrifuged at 20,000 × g for 10 minutes to obtain the supernatant. The supernatant was collected and passed through a combined C18-50/PSA-30 column that had been conditioned with 2 mL of 100% acetonitrile and 2 mL of a 3:1.7:1 (v/v/v) mixture of acetonitrile, methanol, and water. The analytes were eluted with acetonitrile. Both the filtrate and eluate were subsequently applied to an activated C18-30 column conditioned with a solvent of similar polarity to the sample. After passage, the analytes were eluted with 80% acetonitrile. The resulting solution (∼1.5 mL) was mixed thoroughly, and 200 µL was used for analysis.

### UPLC-MS/MS Measurement Conditions

UPLC-MS/MS analysis was performed using ACQUITY UPLC system, which included an ACQUITY UPLC binary pump and a sample manager (Waters, Milford, MA, USA). The analytical column used was CAPCELL PAK INERT ADME-HR (2.0 mm I.D. × 100 mm, particle size 3 μm; Osaka Soda Co., Ltd., Osaka, Japan). The column temperature was maintained at 40 °C, and the sample injection volume was 5 μL. Mobile phase A consisted of 10 mM ammonium formate, and mobile phase B was acetonitrile containing 0.1% formic acid. The flow rate was set to 0.3 mL/min. The mobile phase system started at 99% A for 1 min. A linear gradient was used to ramp from 80% A to 1% A within 5 min and maintained at 1% A for 2 min. The mobile phase was then returned to 99% within 0.01 min and maintained at 99% A for 2 min to equilibrate the column for the next sample.

The MS/MS analyses were conducted on a tandem quadrupole mass spectrometer (ACQUITY TQD; Waters) equipped with an electrospray ionization (ESI) source. Phenobarbital and phenobarbital-d5 were analyzed in the negative ionization mode, while other analytes were analyzed in the positive ionization mode.

Quantification was carried out by selected reaction monitoring (SRM) based on peak area measurements. The instrument parameters were set as follows: capillary voltage, 2.8 kV; source temperature, 150 °C; desolvation temperature, 450 °C; desolvation gas and cone gas (nitrogen) flow rates, 800 and 50 L/h, respectively; and collision gas (argon) flow rate, 0.15 mL/min. All data were acquired in centroid mode and processed using MassLynx NT 4.1 software with the QuanLynx program (Waters).

### Method validation

The method was validated according to the US FDA guidelines on bioanalytical method validation and related documents^15)^. The validation parameters included linearity, intra-day and inter-day precision, matrix effects, and recovery of spiked samples. The intra- and inter-day accuracies and precisions were determined by conducting 3 experiments in the course of a single day and by conducting one experiment on 5 different days, respectively.

Intra-day and inter-day precision were evaluated at three concentration levels within the validated calibration range (low, medium, and high). For each concentration level, three replicate samples were prepared and analyzed, and the measurements were repeated over five consecutive days. Precision was expressed as the coefficient of variation (CV). Accuracy and precision were required to be within ± 20% at the lower limit of quantification (LOQ) and within ± 15% at all other concentration levels.

Matrix effects and recovery were evaluated as follows. For the assessment of matrix effects, whole blood was deproteinized with methanol and acetonitrile, and the resulting supernatant was passed through a mini-cartridge. The eluate obtained by mini-cartridge extraction was then transferred into test tubes containing the analytes and IS that had been previously added and evaporated to dryness under a stream of nitrogen (Aeb). Similarly, purified water was processed in the same manner as whole blood, and the resulting eluate was transferred into test tubes containing the analytes and IS that had been previously added and evaporated to dryness under nitrogen (Ans). Each sample was analyzed, and the matrix effect was calculated from the obtained peak areas using the following equation: matrix effect (%) = (Aeb / Ans) × 100.

Extraction recovery was evaluated by comparing two sets of samples. In one set, whole blood was deproteinized with methanol and acetonitrile, and the resulting supernatant was passed through a mini-cartridge. The eluate was then transferred into test tubes containing the analytes and internal standard (IS) that had been previously added and evaporated to dryness under a stream of nitrogen (Aeb). In the other set, the analytes and IS were added directly to whole blood prior to deproteinization with methanol and acetonitrile, and the resulting supernatant was filtered through a mini-cartridge to obtain the eluate (Aex). Both sets were analyzed, and extraction recovery was calculated from the obtained peak areas using the following equation: recovery (%) = (Aex / Aeb) × 100.

## Results

### Examination of Phospholipid Removal Effect Using Solid-Phase Mini Cartridges

The SRM transitions for phospholipids are shown in Table 1. Total ion chromatograms of the samples obtained under each condition using these SRM transitions are shown in Figure 3. Distinct phospholipid peaks were observed with the centrifugal evaporator method, whereas the peaks were markedly reduced and suppressed when the column method was applied. In particular, pronounced differences were observed for precursor ions at m/z 524, 760, and 786. A comparison of the peak areas corresponding to these SRM transitions is presented in Table 2.

**Table 1.**
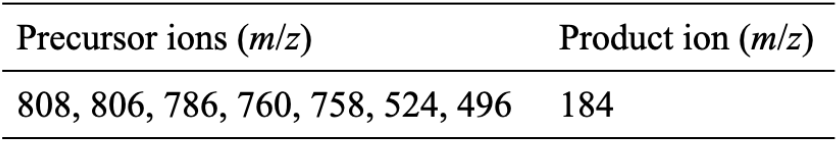
SRM transitions for detection of phospholipids.

**Table 2.**
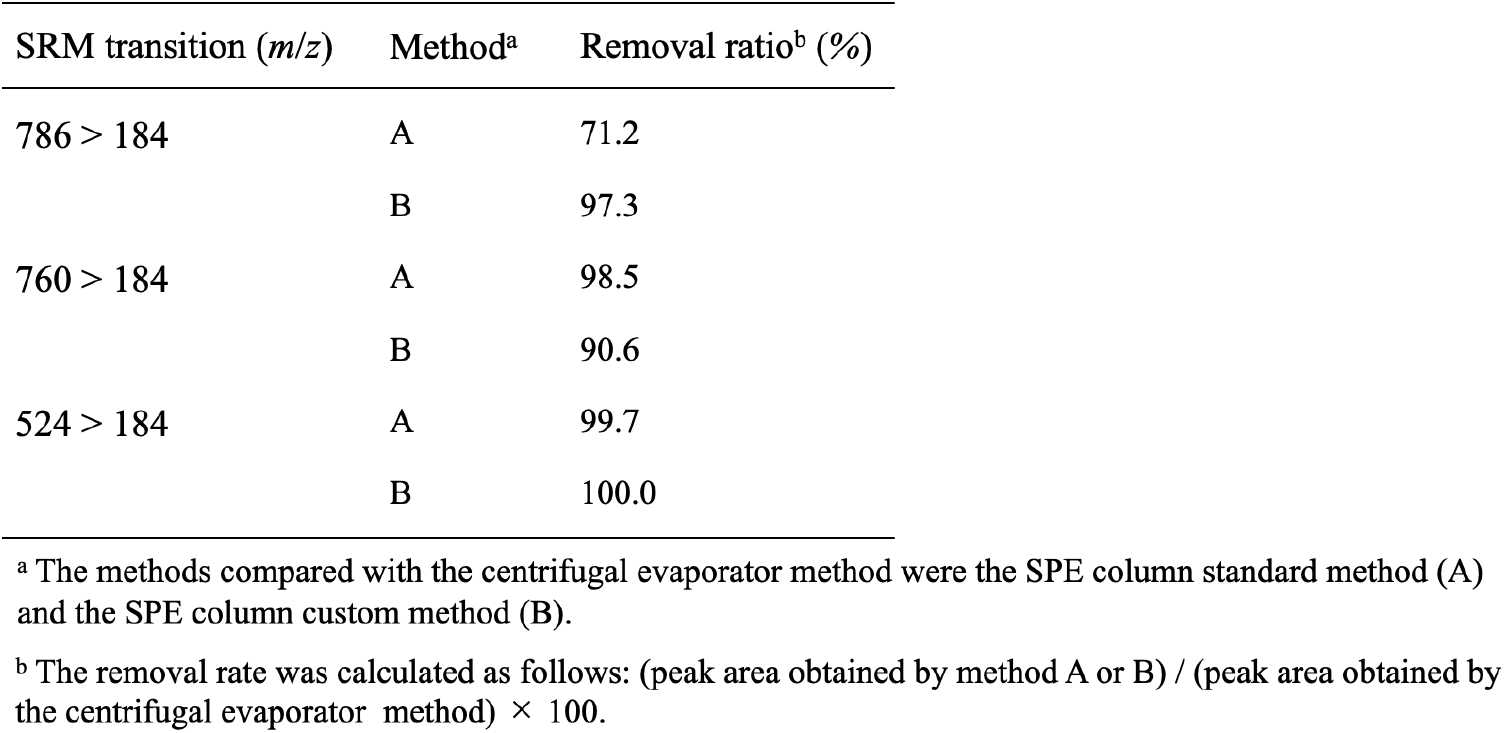
Comparison of peak areas with the centrifugal evaporator method for each SRM transition.

**Figure. 3.**
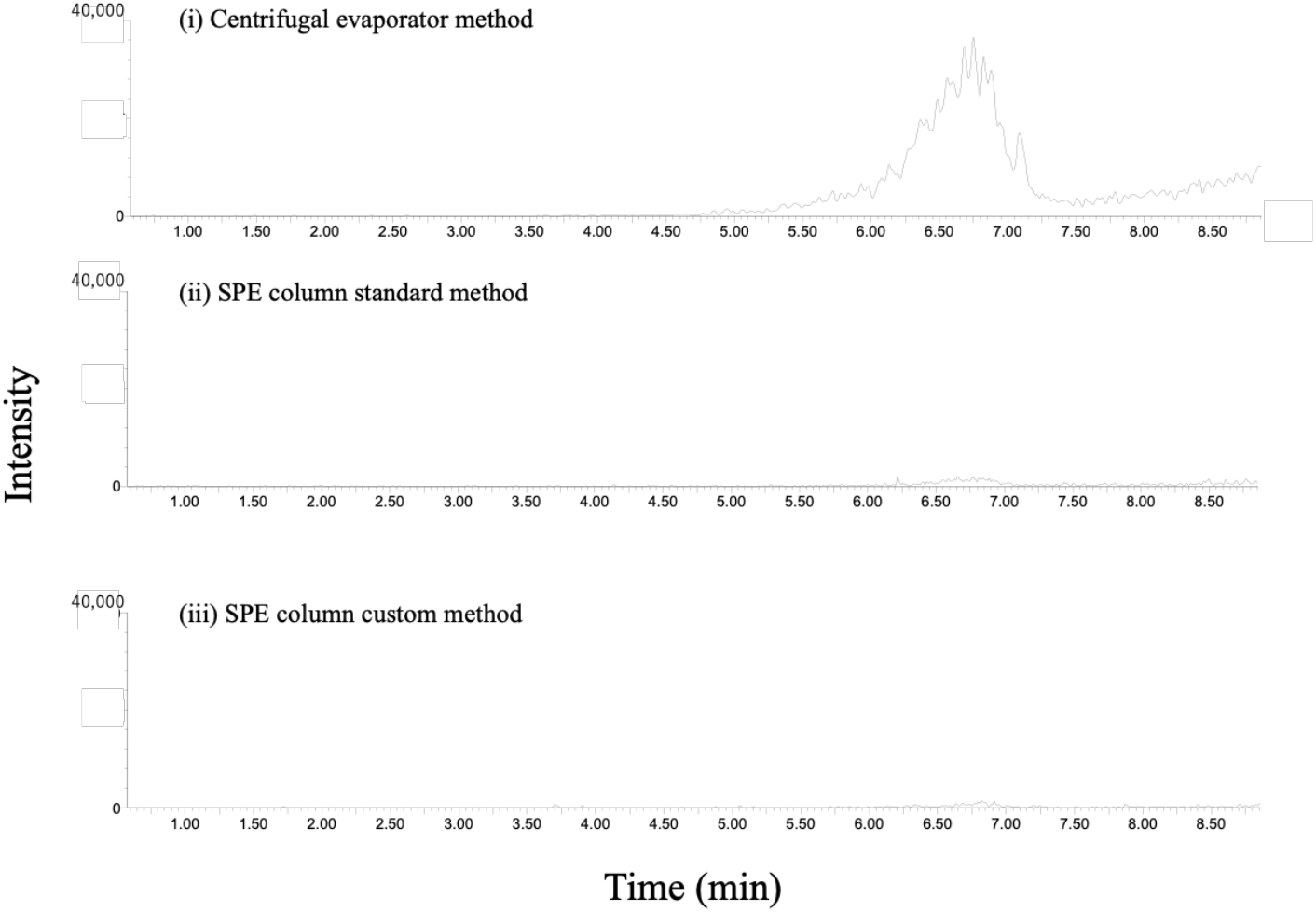
Total ion chromatogram of phospholipids in whole blood samples.

When compared with the peak areas obtained using the centrifugal evaporator method, phospholipid removal rates ranged from 71.2% at the lowest to greater than 90% in other cases. Furthermore, among the column-based methods, the custom column method consistently achieved phospholipid removal rates exceeding 90%, indicating a stable and efficient removal performance. Based on these findings, the custom column method was selected for sample pretreatment.

### Optimization of analytical conditions

The SRM transitions for the target analytes and IS, as well as other MS/MS analytical conditions, are shown in Table 3. The chromatograms of the target substances and IS are shown in Figure 4. Measurements were performed at therapeutic concentrations for all samples. The following concentrations were used: Acetaminophen-d4 at 20 μg/mL, Caffeine-d9 at 10 μg/mL, Phenobarbital-d5 at 10 μg/mL, and Diazepam-d5 at 1 μg/mL. Acetaminophen was detected first at 2.53 minutes, and all substances were detected by 5.11 minutes, when Diazepam-d5 was observed. All peaks were of good quality.

**Table 3.** SRM transition and parameters for detection of the analytes and IS.

| Analyte | Molecular weight | Retention time (min) | SRM transition (*m/z*) | Cone voltage (V) | Collision energy (eV) |
| --- | --- | --- | --- | --- | --- |
| Acetaminofen | 151.16 | 2.53 | 152 > 93 | 26 | 22 |
| Caffeine | 194.19 | 2.64 | 195 > 138 | 40 | 20 |
| Risperidone | 410.49 | 3.49 | 411 > 191 | 50 | 40 |
| Phenobaribital | 232.24 | 3.57 | 231 > 188 | 30 | 10 |
| 7-aminoflunitrazepam | 283.31 | 3.66 | 284 > 135 | 50 | 28 |
| Flunitrazepam | 313.28 | 4.60 | 314 > 268 | 46 | 26 |
| Acetaminofen-*d*<sub>4</sub> (IS) | 155.2 | 2.54 | 156 > 114 | 30 | 16 |
| Caffeine-*d*<sub>9</sub> (IS) | 203.25 | 2.61 | 204 > 144 | 40 | 18 |
| Phenobarbital-*d*<sub>5</sub> (IS) | 237.27 | 3.56 | 236 > 193 | 30 | 10 |
| Diazepam-*d*<sub>5</sub> (IS) | 289.77 | 5.11 | 290 > 154 | 46 | 28 |

### Reliability of the Measurement Method

The results for linearity, measurement range, and limits of detection are summarized in Table 4. For all analytes tested, the correlation coefficients demonstrated excellent linearity, with values exceeding 0.99. Although the therapeutic blood concentrations of caffeine, acetaminophen, and phenobarbital are relatively high, the proposed method covers a wide concentration range extending from therapeutic to lethal levels. In contrast, risperidone and flunitrazepam exhibit therapeutic blood concentrations approximately three orders of magnitude lower than those of caffeine and acetaminophen; nevertheless, good linearity was achieved over a concentration range spanning from below the therapeutic level to lethal concentrations^16)^.

**Table 4.** Regression equation, correlation coefficients (*r*) and limits of detection for the analyzed standards.

| Analyte | Range (μg/mL) | Regression equation <sup>a</sup> | Correlation coefficient (*r*) | LOD (μg/mL) |
| --- | --- | --- | --- | --- |
| Acetaminophen | 0.05-200 | $y = 0.0123x + 0.0172$ | 0.9994 | 0.025 |
| Caffeine | 0.01-200 | $y = 0.0896x + 0.0847$ | 0.9988 | 0.005 |
| Risperidone | 0.0001-2 | $y = 6.4865x - 0.0091$ | 0.9999 | 0.00005 |
| Phenobaribital | 0.5-200 | $y = 0.0704x + 0.1182$ | 0.9998 | 0.25 |
| 7-aminoflunitrazepam | 0.001-2 | $y = 2.4605x + 0.0238$ | 0.9997 | 0.0005 |
| Flunitrazepam | 0.0005-2 | $y = 1.9363x + 0.0012$ | 0.9999 | 0.00025 |

<sup>a</sup> Values are the means of five replicate determinations.

The intra- and inter-day accuracies and precisions were evaluated at four levels, the results of which are summarized in Table 5. The intra-day accuracies and precisions were 98.1 – 110.9% and 0.2 – 17.4%, respectively. The inter-day accuracies and precisions were 97.8 – 112.4% and 0.5 – 9.2%, respectively. Since accuracy values closer to 100% indicate better performance, and lower precision values closer to 0% are preferable, slightly higher precision values were observed at the lower limits of quantification for caffeine and risperidone. Nevertheless, overall intra-day and inter-day reproducibility demonstrated generally acceptable performance.

**Table 5.**
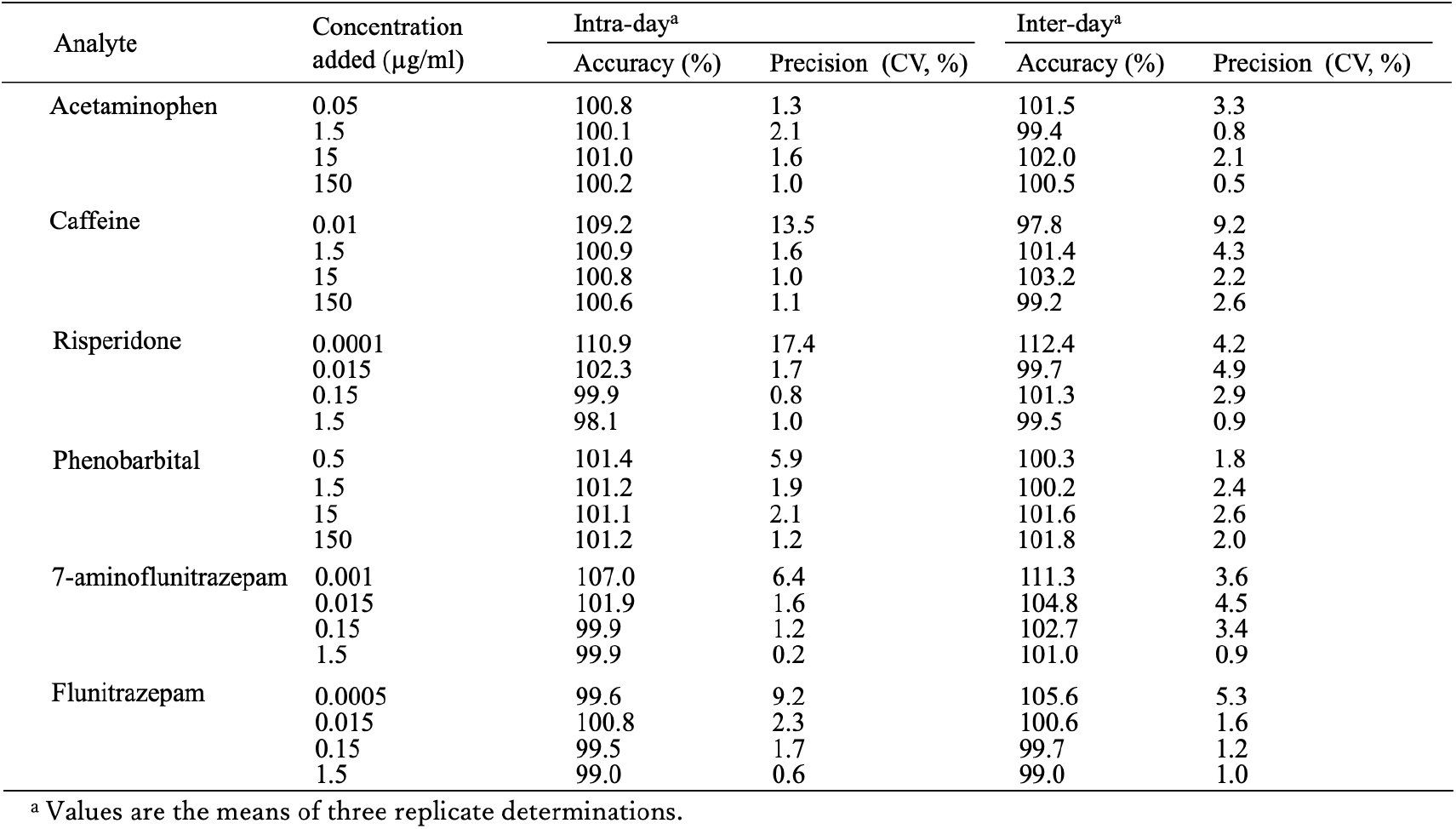
Intra-day and inter-day accuracy and precision data of the analytes in whole blood.

The recovery efficiencies and the matrix effects of the samples are shown in Table 6. The recovery efficiencies were in the range of 72.9 – 95.1%. Matrix effects were observed ranging from 91.2% – 113.0%. The recovery efficiencies closer to 100% indicates better performance, and a matrix effect closer to 100% suggests minimal influence. Therefore, both parameters demonstrated acceptable performance for the proposed method.

**Table 6.**
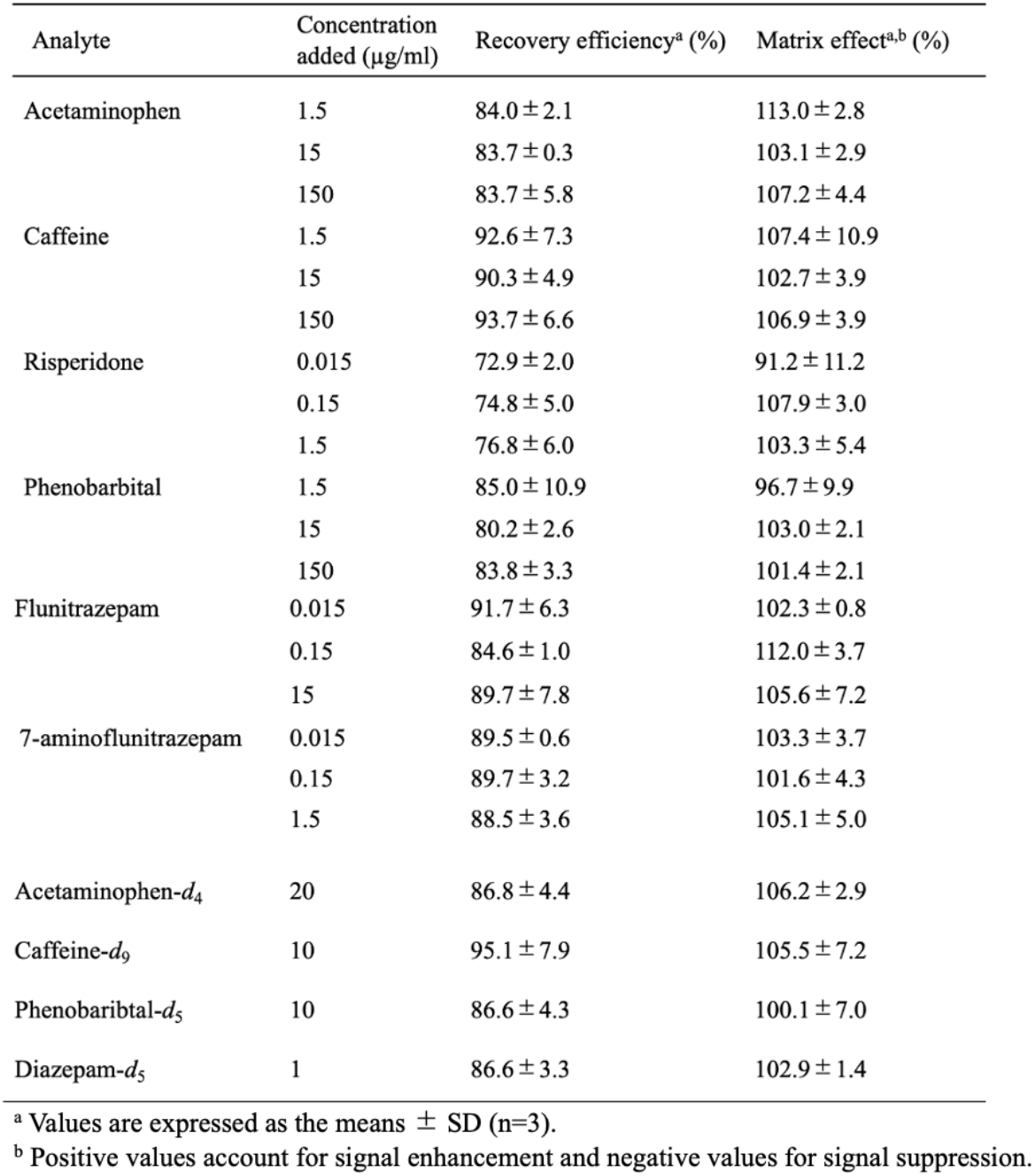
The recovery efficiencies and matrix effects of the analytes in whole blood.

| Analyte | Concentration added ($\mu\text{g/ml}$) | Recovery efficiency<sup>a</sup> (%) | Matrix effect<sup>a,b</sup> (%) |
| --- | --- | --- | --- |
| Acetaminophen | 1.5 | 84.0 ± 2.1 | 113.0 ± 2.8 |
| | 15 | 83.7 ± 0.3 | 103.1 ± 2.9 |
| | 150 | 83.7 ± 5.8 | 107.2 ± 4.4 |
| Caffeine | 1.5 | 92.6 ± 7.3 | 107.4 ± 10.9 |
| | 15 | 90.3 ± 4.9 | 102.7 ± 3.9 |
| | 150 | 93.7 ± 6.6 | 106.9 ± 3.9 |
| Risperidone | 0.015 | 72.9 ± 2.0 | 91.2 ± 11.2 |
| | 0.15 | 74.8 ± 5.0 | 107.9 ± 3.0 |
| | 1.5 | 76.8 ± 6.0 | 103.3 ± 5.4 |
| Phenobarbital | 1.5 | 85.0 ± 10.9 | 96.7 ± 9.9 |
| | 15 | 80.2 ± 2.6 | 103.0 ± 2.1 |
| | 150 | 83.8 ± 3.3 | 101.4 ± 2.1 |
| Flunitrazepam | 0.015 | 91.7 ± 6.3 | 102.3 ± 0.8 |
| | 0.15 | 84.6 ± 1.0 | 112.0 ± 3.7 |
| | 15 | 89.7 ± 7.8 | 105.6 ± 7.2 |
| 7-aminoflunitrazepam | 0.015 | 89.5 ± 0.6 | 103.3 ± 3.7 |
| | 0.15 | 89.7 ± 3.2 | 101.6 ± 4.3 |
| | 1.5 | 88.5 ± 3.6 | 105.1 ± 5.0 |
| Acetaminophen-*d*<sub>4</sub> | 20 | 86.8 ± 4.4 | 106.2 ± 2.9 |
| Caffeine-*d*<sub>9</sub> | 10 | 95.1 ± 7.9 | 105.5 ± 7.2 |
| Phenobarbital-*d*<sub>5</sub> | 10 | 86.6 ± 4.3 | 100.1 ± 7.0 |
| Diazepam-*d*<sub>5</sub> | 1 | 86.6 ± 3.3 | 102.9 ± 1.4 |

<sup>a</sup> Values are expressed as the means ± SD (n=3).
<sup>b</sup> Positive values account for signal enhancement and negative values for signal suppression.

## Discussion

By utilizing the Smart-SPE solid-phase extraction column from AiSTI Science Co., Ltd., it was confirmed through chromatography that the removal of phospholipids, which could not be completely eliminated by methanol and acetonitrile-based deproteinization, was effectively achieved. Additionally, the minimal matrix effects observed indicate that Smart-SPE has a high efficacy in removing impurities. Since no washing step was employed, the sensitivity for highly polar compounds such as acetaminophen and caffeine were sufficiently maintained. Furthermore, even for drugs with low therapeutic concentrations, such as flunitrazepam and risperidone, adequate sensitivity was obtained.

Although analytical methods for drug poisoning are being actively investigated worldwide^17-19)^, many existing approaches primarily target prescribed drugs, such as psychotropic agents, whereas substances present at higher blood concentrations, including caffeine, are often analyzed using separate methods^9),11-12), 20)^. The present study offers a distinct advantage in that it enables the simultaneous quantification of a wide range of drugs with markedly different blood concentration levels, including OTC medications. This capability eliminates the need for prior sample dilution and allows more rapid clinical feedback for substances such as caffeine and acetaminophen, which have traditionally required separate analytical procedures.

Because intra-day and inter-day accuracies and precisions, recovery efficiencies, and matrix effects were all generally acceptable, it was suggested that this measurement method has high practicality as a technique for measuring blood drug concentrations, including OTC medications. Based on these results, the present method using Smart-SPE is considered to be sufficiently practical as a novel sample pretreatment approach for the measurement of blood drug concentrations.

## Conclusion

In this study, we investigated a simultaneous quantitative method for blood drugs using UPLC-MS/MS with solid-phase mini-cartridge Smart-SPE as a pretreatment method. The developed measurement method was demonstrated to be sufficiently applicable for testing in emergency medicine and forensic fields related to poisoning cases.

## Data Availability

All data produced in the present study are available upon reasonable request to the authors

## References

1. Cabinet Office: Current Status of Over-the-Counter Drug Abuse and Dependence. [Online]. Available from: URL: https://www8.cao.go.jp/kisei-kaikaku/kisei/meeting/wg/2310_04medical/231116/medical04_02.pdf

2. Masayuki Hirose, Akihiko Hirakawa, Wakana Niwa, et al.: Acute Drug Poisoning among Adolescents Using Over-the-counter Drugs: Current Status. Journal of Pharmaceutical Sciences, 141:1389–1392, 2021

3. Takanao Hashimoto, Yudai Kaneda, Akihiko Ozaki, et al.: Eleven-Year Trend of Drug and Chemical Substance Overdose at a Local Emergency Hospital in Japan. Cureus, 14(12), 2022

4. Wakako Hikiji, Yasuyuki Okumura, Toshihiko Matsumoto, et al.: Identification of Psychotropic Drugs Attributed to Fatal Overdose—A Case-control Study Using Data from the Tokyo Medical Examiner’s Office and Prescriptions. Seishin Shinkeigaku Zasshi, 118(1):3–13, 2016

5. Yuichi Uwai, Tomohiro Nabekura: Surveillance of Drug Overdose and Identification of Its Risk Factors Using Multivariate Analysis of the Japanese Adverse Drug Event Report Database. Asian Journal of Psychiatry, 65, 2021

6. W Victor R Vieweg, Mehrul Hasnain, Jules C Hancox, et al.: Risperidone, QTc Interval Prolongation, and Torsade de Pointes: A Systematic Review of Case Reports. Psychopharmacology (Berlin), 228(4):515–24, 2013

7. Takafumi Nakano, Yoshihiko Nakamura, Keisuke Sato, et al.: Adrenaline-Resistant Anaphylactic Shock Caused by Contrast Medium in a Patient After Risperidone Overdose: A Case Report. Journal of Pharmaceutical Health Care Sciences, 9(1):23, 2023

8. Masayuki Hirose: Responding Calmly to Poisoning Cases Encountered During Night Shifts—Common Symptoms and Medications Used. Yakujii, 64:44–48, 2022

9. Kiyotaka Usui, Yuji Fujita, Yoshito Kamijo et al.: LC-MS/MS method for rapid and accurate detection of caffeine in a suspected overdose case. J Phamacol Toxicol Methods. 107, 2021

10. Dylan Mantinieks, Jennifer Schumann, Olaf H Drummer et al.: Stimulant use in suicides: A systematic review. Forensic Sci Int. 338, 2022

11. Chrisyian C Toquica Gahona, Ashwin Kodagnur Bharadwaj, Monarch Shah et al.: Treatment of Lethal Caffeine Overdose with Haemodialysis.: A Case Report and Review. J Crit Med(Targu Mures). 8(4): 279–287, 2022

12. Simone Cappelletti, Daria Piacentino, Vittorio Fineschi et al.: Caffeine-Related Deaths: Manner of Deaths and Categories at Risk. Nutrients. 10(5): 611, 2018

13. Shoichi Imakawa, Tatsuhiro Kuwahara, Kazuki Nagashima et al.: Status and issues of instrumental analysis for toxic substances in emergency intensive care unit. Journal of the Japanese Society of Emergency Medicine, 23: 711–716, 2020.

14. Takayoshi Suzuki, Tadashi Ogawa, Masae Iwai et al.: Simultaneous quantification of multiple antidiabetic drugs in whole blood by ultra-performance liquid chromatography-tandem mass spectrometry. Medical Mass Spectrometry, 7(1): 35–42, 2023.

15. US FDA: Guidance for Industry: Bioanalytical Method Validation, 2018. Available from: URL: https://www.fda.gov/downloads/drugs/guidances/ucm070107.pdf

16. Martin Schulz, Stefanie Iwersen-Bergmann, Hilke Andresen et al.: Therapeutic and toxic blood concentrations of nearly 1,000 drugs and other xenobiotics. Critical Care. 16: R136, 2012

17. Chi-Wei Lee, Hung Su, You-Da Cai et al.: Rapid Identification of Psychoactive Drugs in Drained Gastric Lavage Fluid and Whole Blood Specimens of Drug Overdose Patients Using Ambient Mass Spectrometry. Mass Spectrom(Tokyo). 6(2), 2017

18. A Lopez-Rabunal, E Lendoiro, M Comcheiro et al.: LC-MS-MS Method for the Determination of Antidepressants and Benzodiazepines in Meconium. J Anal Toxicol. 44(6): 580–588, 2020

19. Emily Elenstal, Henrik Green, Robert Kronstrand et al.: Intralipid as a matrix additive for evaluating hyperlipidemic postmortem blood. J Anal Toxicol. 47(6): 529–534, 2023

20. Hiroyasu Maise, Ayano Tanaka, Takayuki Shinoo, et al.: Simultaneous measurement of concentrations of caffeine and ibuprofen in blood by LC-MS/MS and pharmacokinetic analysis after oral intake of canned coffee. Medical Testing, 64(4): 413–420, 2015.

